# Co-designed Sleep Health Program Improves Sleep Health of Australian First Nations Adolescents: Findings from a Pilot Study

**DOI:** 10.64898/2026.06.11.26355484

**Authors:** Yaqoot Fatima, Roslyn Von Senden, M Mamun Huda, Aunty Joan Marshall, Daniel P Sullivan, Romola S. Bucks, Azhar Hussain Potia, Simon S. Smith, Sarah Blunden, Lisa McDaid, Markesh Fanti, Peter R. Eastwood, Stephanie Yiallourou, Jen Walsh, Abdullah Mamun, Stephanie King, Sharon Varela, Shaun Solomon, Timothy C Skinner

## Abstract

**Background:** Adolescent sleep health is a growing public health concern, yet no culturally responsive sleep health program has been developed for Aboriginal and Torres Strait Islander (First Nations) young people. This study reports the outcomes and acceptability of Australia’s first co-designed sleep health program for First Nations adolescents.

**Methods:** The Let’s Yarn About Sleep adolescent program was co-designed with First Nations community members from 23 Traditional groups, involving 174 Elders, adolescents, parents, carers, and service providers. The program drew on an Aboriginal pedagogical framework and the COM-B behaviour change model, integrating Western and First Nations sleep science, and was delivered by Aboriginal Youth Workers trained as Sleep Coaches. Outcomes included self-reported improvements in sleep knowledge, sleep timing and continuity, sleep quality, overall sleep health, and psychological distress. Post-program changes in outcomes were assessed using linear mixed-effects regression analyses. Program ratings and yarning-based feedback assessed acceptability.

**Findings:** 70 First Nations young people participated in the program (median age 13.0 years, range 12–18; 67.1% female). Sleep knowledge improved substantially, with the mean composite score increasing from −0.65 (SD 1.23) at baseline to 0.82 (SD 1.27) at follow-up, a large effect (Cohen’s *d* = 1.18; *p* < 0.001). A significant improvement was observed in the overall sleep health score, representing a medium effect (Cohen’s *d* = 0.63; β = 0.68, 95% CI: 0.31–1.04; *p* < 0.001). Psychological distress showed a directional reduction that did not reach statistical significance (Cohen’s *d* = 0.32; β = −0.49, 95% CI: −1.08–0.09; *p* = 0.097), though a modest beneficial effect cannot be excluded. High acceptability was reflected in program ratings and qualitative feedback, with participants reporting greater sleep awareness, improved sleep behaviours, and strong community engagement with the program.

**Interpretation:** A culturally grounded, co-designed sleep health program can improve sleep knowledge and overall sleep health and achieve high acceptability among First Nations adolescents. Community leadership, local delivery, and the embedding of First Nations worldviews are likely central to achieving impact, highlighting a promising pathway to address sleep health inequities

**Funding:** Medical Research Future Fund (APP1201569). No competing interests.

**Research in context:** *Evidence before this study:* We searched PubMed, Scopus, and PsycINFO for peer-reviewed studies published between January 2010 and Sep 2025 with language restrictions, using the search terms: *“adolescent sleep health”*, *“sleep education”*, *“Indigenous”*, *“First Nations”*, *“co-design”*, and *“community-led delivery”*. Reference lists of relevant reviews were also screened. Evidence from systematic reviews and meta-analyses showed that most adolescent sleep interventions are delivered in school or community settings and are grounded in behavioural or cognitive–behavioural approaches. The existing sleep health programs are mainly delivered by sleep health researchers or healthcare providers in school settings over short durations (four to six weeks) and have produced modest short-term improvements in some aspects of sleep health. None of these programs was co-designed or culturally adapted for First Nations communities or offered in community settings. Further, most tools used to assess sleep health and knowledge were developed in Western contexts, with little consideration of First Nations’ conceptualisation of sleep health or of cultural and contextual factors that impact it. No prior studies have integrated First Nations governance and culturally grounded learning frameworks to improve the sleep health of First Nations adolescents.

*Added value of this study:* This is Australia’s first sleep health program for First Nations adolescents. This co-designed program combined Western and First Nations sleep science and privileged First Nations cultural knowledge and practices to address the sleep health needs of First Nations adolescents holistically. This community-based program demonstrated significant improvements in sleep knowledge and sleep health, with strong community engagement and satisfaction.

*Implications of all the available evidence:* Co-designed and community-led approaches can help improve the sleep health of First Nations young people. Embedding First Nations leadership, local workforce development, and community governance structures into sleep health programs could improve program engagement and acceptability and potentially lead to improvements in sleep health. Future research should focus on integrating culturally responsive sleep health programs within school and community well-being systems to reduce sleep health inequities and improve life outcomes for First Nations adolescents.

## Introduction

Humans spend approximately one-third of their lives in various states of sleep.1 Both sufficient quantity and optimal quality of sleep are critical for maintaining physical health and social-emotional well-being.2 Evidence from systematic reviews and meta-analyses consistently shows that individuals who have shorter sleep compared to their age group norms are more likely to develop a range of chronic physical and mental health conditions, such as diabetes, obesity, and depression.3,4 Even a single night of short, disturbed sleep results in dysregulation of the endocrine, nervous, and immune systems and reduces our capacity for advanced cognitive activities and emotion regulation.5

Although healthy sleep matters across the lifespan, adolescence warrants particular attention given its developmental importance and heightened vulnerability to poor sleep.^6,7^ Pubertal hormonal changes shift the circadian clock forward by one to two hours, delaying evening sleepiness,^8^ while shifting social patterns, stimulating technology use, and growing self-regulatory demands further undermine sleep quality.^9^ As a result, many adolescents experience chronic sleep restriction and irregular sleep, with broad consequences for psychological, physical, and social health that can shape long-term life trajectories.^10^

Despite the established role of good sleep, most Australian adolescents are not getting the sleep they need. Data from the Longitudinal Study of Australian Children show that only one in four young people aged 12–15 and around half of those aged 16–17 meet the recommended sleep guidelines.^11^ Aboriginal and Torres Strait Islander (hereafter referred to as First Nations) adolescents face an even steeper challenge. Emerging evidence indicates they are at greater risk of short, interrupted, and poor-quality sleep, with downstream effects on health and well-being.^12^

Sleep health inequities faced by First Nations peoples are rooted in colonisation, ongoing structural disadvantage, and broader social determinants of sleep, such as neighbourhood disadvantage, exposure to light and noise pollution, socio-economic status, and access to safe, stable housing.^13^ Importantly, these factors operate at multiple levels: macro (e.g., policy, structural inequality), meso (community, environment), and micro (household, individual). The cumulative impact of these social and environmental conditions shapes young people’s knowledge, attitudes, capacities, and opportunities to improve their sleep health.

At the same time, First Nations communities have long understood the importance of sleep. Yolŋu Elders from Arnhem Land and their communities describe sleep as integral to health and well-being within a holistic Social and Emotional Wellbeing (SEWB) framework that encompasses spirituality, kinship, and connection to land and Country.^14^ This understanding is echoed in research with First Nations peoples from other communities in Australia, where sleep is highlighted as more than rest and restoration and a key aspect of spiritual well-being^15^ and the impact of sleep health can extend to an individual’s connection to Country, kinship, culture, spirituality and ancestry.^16^

Regrettably, the evidence base on sleep health of First Nations peoples remains thin, and no program has ever been developed to support Australian First Nations young people in improving their sleep. In non-First Nations adolescent populations, sleep-health interventions have generally been delivered through school- or community-based settings using structured group formats led by trained educators or a healthcare team.^17,18^ These programs typically combine psycho-education with behavioural components such as sleep hygiene training, stimulus control, and cognitive restructuring and have shown moderate improvement in sleep knowledge.^19^ However, mainstream health programs do not capture cultural and contextual considerations shaping sleep health and often fail to resonate with First Nations communities.^20^ To advance sleep health equity in First Nations communities, a whole-of-community approach, co-designed with communities, is needed to ensure cultural responsiveness, contextual relevance, and participant empowerment for meaningful change in sleep health.

Recognising the need for a community-led sleep health program, we sought to co-develop one that used two-way knowledge processes, privileging First Nations worldviews and both First Nations and Western sleep sciences. Here, we report the program’s delivery and outcomes, examining its acceptability and effectiveness in improving sleep health and psychological well-being among First Nations adolescents.

## Methods

### Project setting

This project was conducted in Mount Isa, a remote town in North West Queensland. Approximately a quarter of the population identifies as First Nations Australian (22%), compared to 4.6% across Queensland and 3.2% across the nation; 13.6% of the town’s population is aged 10-19 years.^21^

### Study design

The study used a mixed-methods, pre–post design to assess changes in sleep health knowledge, overall sleep health, and psychological distress from program start to completion. Full details of the study design and measurement approaches have been described in the published protocol paper.^22^

### Program co-design framework and governance

The program co-design process followed a strengths-based (focusing on existing community assets, capabilities, and cultural knowledge) and knowledge co-creation approach ^23^. The theoretical framework informing the program integrated Yunkaporta’s Eight Ways of Learning framework for culturally responsive program delivery ^24^ and the COM-B model (Capability, Opportunity, Motivation, and Behaviour) to guide behavioural change.^25^

The Eight Ways of Learning framework is a First Nations pedagogical approach that grounds teaching and learning in First Nations ways of knowing, including storytelling, place-based learning, and community relationships to ensure culturally responsive program delivery. This framework privileges the eight interconnected ways Aboriginal communities have always taught and shared knowledge through story, symbol, movement, place, deconstruction, community relationships, and non-linear thinking. We applied this framework to ensure that knowledge is shared in ways informed by First Nations ways of knowing, being, and doing.

The COM-B framework informed the inclusion of strategies that enhanced participants’ capability (sleep education, skill-building), physical and social opportunities (peer and family support, cultural practices), and reflective motivation (goal setting, self-monitoring).

At the outset, a Community Steering group of First Nations Elders was established to guide all aspects of the program’s development, delivery, evaluation, and dissemination, ensuring that processes and outcomes reflected First Nations worldviews and community priorities.

### Co-design of sleep health program and study measures

Let’s Yarn About Sleep (adolescent sleep health program) was co-designed with 174 stakeholders, including Elders, young people, parents and carers from 23 Traditional groups, and service providers from schools, youth, housing, and health services. The co-design process was guided by a Community Steering Group and undertaken over 18 months across three phases: learning, designing, and refining. The learning phase focused on understanding community perspectives and priorities and co-defining the problem from First Nations young people’s perspectives; the designing phase involved developing the program and its resources; and the refining phase focused on further iteration and refinement with participants.^26^

Co-design workshops were conducted with three key groups, with each workshop lasting approximately 90–120 minutes. Six workshops were held with young people (*n* = 44), with a median age of 14.0 years (range 12–18); 54.5% were female. Ten workshops were conducted with service providers (*n* = 27), with a median age of 35.0 years (range 19–58); 85.2% were female. A further ten workshops were held with parents, carers, and Elders (*n* = 96), with a median age of 51.5 years (range 19–83); 74.7% were female.

The co-design process used culturally grounded methods, including storytelling, painting, and yarning. These approaches supported participants in sharing their perspectives on sleep and identifying what constitutes an effective and sustainable program. Insights from this process informed the program’s content, structure, and delivery, ensuring it reflects community priorities, local knowledge systems, and First Nations perspectives on holistic well-being.

At the outset, the Community Steering Group reviewed standard validated tools commonly used in adolescent sleep research to assess their usability and cultural relevance. These included measures of sleep-related knowledge, beliefs, and attitudes ^27^, sleep-facilitating and sleep-inhibiting practices ^28^, sleep difficulties ^29^ and sleep quality.^30^ The group advised that these tools may not reflect the lived realities of First Nations young people and may not adequately capture sleep health from a First Nations perspective. They highlighted the need to develop tools that are specific to First Nations contexts and worldviews. In response, the co-design process also focused on developing culturally responsive tools, including a sleep health assessment and a sleep diary (available on program website: https://letsyarnaboutsleep.org/resources/research), grounded in local understandings of sleep and well-being within a strengths-based SEWB framework.

The final program, along with the sleep health assessment tool and sleep diary, was endorsed by the Community Steering Group. Detailed co-design methods will be reported in a separate publication.

### Overview of the co-designed sleep health program

The final program comprised pre- and post-program data collection and five group-based sessions delivered over ten weeks in local communities. Local Aboriginal Youth Workers were trained as Sleep Coaches to deliver the program. Each session lasted approximately 90–120 minutes and began with an Acknowledgment of Country, recognising Traditional Custodians of the land and connecting participants to the place. Sessions concluded with a Dreamtime Story, reinforcing cultural values and community connection. The Sleep Coaches maintained regular contact with participants and families through phone calls, home visits, and community check-ins to support continuity and reinforce learning between sessions (Table 1).

**Table 1:** Summary of Let’s Yarn About Sleep (adolescent sleep health program) sessions and activities.

| Session | Aim | Learning Outcomes | Key Activities | Setting |
| --- | --- | --- | --- | --- |
| 1. Yarn About Sleep | To explore the importance of sleep and its role in overall health and well-being | Participants understood basic sleep physiology, identified personal and community sleep protectors and disruptors, and set individual sleep goals. | Sleep Routine Check, Hula Hoop Activity (sleep protectors and disruptors at self, family, community levels), Persona Identification, introduction to sleep hygiene | Community, schools or youth hub. |
| 2. Deadly Sleep: What Is It and Why Does It Matter? | To understand the impact of sleep loss and introduce sleep hygiene practices. | Participants recognised links between sleep, physical and mental health, and identified healthy sleep habits. | Word-for-Emotion Activity, Feltman® and Animal Cards, sleep hygiene education | Community, schools or youth hub. |
| 3. Deadly Sleep: How Do I Get It? | To build skills in managing thoughts and emotions that affect sleep. | Participants learned relaxation and cognitive strategies to improve sleep quality. | Sleep hygiene education, worry tree and gratitude tree, sleep affirmations. | Community, schools or youth hub. |
| 4. Connecting with Country | To strengthen the connection with Country and cultural identity in relation to sleep health | Participants deepened their cultural understanding of traditional sleep practices and their connection to nature. | Walk on Country with Elders to receive information on bush food, bush medicines, and traditional practices supporting healthy sleep, native seed activities | Outdoors or on Country. |
| 5. Sleep for Strong Souls | To promote holistic well-being of mind, body, and spirit through culturally grounded sleep practices. | Participants practised mindfulness, deep listening, and First Nations relaxation techniques to support healthy sleep | Deep listening, breathing and visualisation exercises. | Outdoors or natural space (e.g., bush or open community area). |
| Graduation Ceremony | To celebrate participant achievements and reinforce ongoing commitment to healthy sleep practices. | Participants reflected on their learning, shared experiences, and celebrated personal and group growth. | Certificate presentation, group reflection, and light refreshments. | Community or school setting. |

### Program promotion and participant recruitment

The program was promoted through community channels, including social media, local radio, primary care networks, youth well-being services, and schools. Sleep Coaches facilitated contact and engagement with First Nations adolescents, parents, and community Elders, and liaised with local youth services to support recruitment and retention.

Eligible participants were First Nations adolescents aged 12–18 who self-reported poor sleep, operationalised as sleeping less than 7 hours per night, difficulty initiating or maintaining sleep, early-morning awakening, or persistent daytime fatigue and sleepiness over the preceding 4 weeks, and/or mental health concerns. Exclusion criteria included clinically diagnosed severe obesity (BMI≥40kg.m^-2^), neuromuscular or cognitive impairment, and chronic lung disease.^31^ All participants and their parents or guardians provided written informed consent before participation. Participant engagement and retention were prioritised through relationship building, regular communication, and compensation (a $50 Gift voucher) as a token of appreciation for their time and contributions.

### Program evaluation

This study used a mixed-methods pre-post (10-week) design to assess program outcomes and acceptability. The primary outcomes of this study were improvement in sleep knowledge, sleep quality and overall sleep health. Secondary outcomes were the reduction in psychological distress scores. For program acceptability and engagement, participants’ ratings of program sessions and post-program yarning with participants and their families were used. Together, these measures allow us to assess whether the program changed what young people knew about sleep, how they slept, how they felt, and whether it was meaningful and acceptable to them and their families.

### Sleep-related outcomes

The following multi-method approach to sleep assessment was deliberately chosen because no single measure can capture the full picture of sleep health, particularly in a community where sleep is understood holistically rather than reduced to a single metric. Furthermore, this approach is consistent with contemporary sleep health frameworks that treat sleep as a multidimensional construct ^32^ and with adolescent sleep research recommending multi-method designs.

### Individual sleep parameters

sleep parameters were assessed across three modalities: questionnaire, sleep diary (seven nights), and actigraphy (seven nights), and are reported in full in Supplementary Table S2. Briefly, these covered sleep timing (bedtime, wake time), sleep duration, sleep onset latency, sleep quality, daytime sleepiness, napping, sleep regularity (operationalised as the within-person standard deviation of bedtime and wake time across nights), sleep concerns and daytime functioning. Objective parameters were derived from wrist actigraphy worn across seven consecutive nights at baseline and follow-upActigraphy data were visually inspected for each participant, with sleep windows manually marked, and sleep parameters (supplementary table) derived using GGIR (https://doi.org/10.5281/zenodo.1051064) within the Cicada software application (https://cicada-actigraphy-suite.readthedocs.io/en/latest/).

### Sleep health knowledge (questionnaire-derived)

A six-item tool assessed participants’ knowledge of recommended sleep duration for their age group, understanding of what happens during sleep, awareness of healthy sleep habits, knowledge of the short- and long-term effects of poor sleep, and understanding of sleep debt and weekend recovery sleep. A composite sleep knowledge score was derived from these six items.

### Overall sleep health (questionnaire derived)

A 14-item sleep health questionnaire at baseline and follow-up, asking them to reflect on their sleep over the preceding two weeks. The questionnaire captured self-reported sleep timing on weekdays and weekends, sleep onset latency, number of nocturnal awakenings, and daytime napping. It also assessed difficulty falling asleep, difficulty staying asleep, feeling unrefreshed on waking, and daytime sleepiness, as well as the perceived functional impact of sleep problems on daily life and the degree of worry about sleep. Together, these items reflect the participant’s own holistic evaluation of their sleep. A composite sleep health score was derived from all 14 items.

### Psychological distress

Participants completed the Strong Souls questionnaire at baseline and follow-up. Strong Souls is a culturally validated measure of SEWB developed specifically for First Nations people.^33^ The 11-item abridged version assessed mood and feelings. A composite psychological distress score was derived from 11 items.

### Derivation and interpretation of sleep-related composite scores

To summarise sleep and psychological well-being outcomes into a single interpretable score, composite variables were derived using Polychoric Principal Component Analysis (pPCA). pPCA reduces a set of related ordinal or mixed-type data items to a single summary score that captures the maximum shared variance among them, making it well-suited to multidimensional constructs such as overall sleep health, where no single item tells the whole story. pPCA was conducted on standardised variables, with individual component scores calculated from the first principal component loadings. Because variables were standardised, all scores are mean-centred around zero; a score of zero represents the sample average, positive values indicate above-average performance, and negative values indicate below-average scores relative to the sample at that time point. This is a mathematically expected feature of pPCA-derived scores, not an indication of an absence of the construct being measured. For sleep knowledge, sleep timing and continuity (subjective and objective), sleep quality and sleep health, higher scores reflect better overall sleep outcomes. For psychological distress, higher scores reflect greater distress.

### Covariates

To account for factors known to independently influence sleep health in adolescents, the following covariates were included in all adjusted regression models and captured at both baseline and follow-up. Socio-demographic covariates included age, sex and primary language spoken at home. Sleep environment covariates included the number of people in the home, the number of rooms, having one’s own bedroom, access to a safe sleeping space, access to a clean sleeping space, neighbourhood environment, general health status, and going to sleep hungry. Behavioural covariates included daily screen time, consumption of caffeinated beverages, and physical activity levels. These covariates were selected a priori based on their theoretical relevance to adolescent sleep health and their established associations with sleep outcomes in the broader literature.

### Program acceptability assessment

Program acceptability was assessed through two complementary approaches. At the end of each session, participants completed a brief session rating to capture their immediate experience of the program, including engagement, delivery, and content. Responses were recorded on a three-point scale: “Deadly” (excellent), “Okay”, and “Needs improvement”. At program completion, written feedback was sought from the participants and yarning sessions were conducted with their parents and carers to gather in-depth qualitative feedback. These conversations were facilitated by First Nations Sleep Coaches in community settings and explored participants’ experiences with the program, whether it met their cultural needs, its acceptability and relevance to their community, whether it should be offered to other First Nations communities, and suggestions for improvement. Attendance and participation in session activities were also recorded throughout the program as additional indicators of engagement and acceptability

### Data analysis

A descriptive analysis was conducted to capture participants’ baseline characteristics and provide a clear demographic profile. Depending on the variable type, we used chi-square tests or t-tests (with non-parametric alternatives where applicable) to compare the outcome variables between pre- and post-program.

The change in composite scores for sleep knowledge, sleep health and psychological distress was determined from pre- to post-program, and regression models adjusting for potential confounding factors were used to calculate effect sizes. Changes in individual sleep health items across the program were also evaluated and reported in Supplementary Table S2. Initially, simple regression models were fitted considering only outcome and time (pre-program vs post-program) variables. Subsequently, adjusted regression models were fitted by incorporating potential covariates. Potential confounders were initially selected a priori based on theoretical relevance and prior evidence. This included age, gender, number of people at home, having one’s own bedroom, neighbourhood environment, general health status, and going to sleep hungry. In addition, bivariate analyses were conducted, and variables associated with the outcome at p<0.20 were considered for inclusion in the final multivariable models. Cluster-robust standard errors were applied in regression models for the survey and actigraphy data (two time points) to account for within-subject correlation. For the sleep diary data, which also included repeated daily measurements (days 1–7), linear mixed-effects models were used to account for the nested structure of days within individuals.

Missing data on covariates were addressed using multiple imputation (MI) techniques via chained equations (100 iterations; 30 imputed datasets), with imputation validity verified by comparing covariate distributions before and after imputation. We tested for Missing Completely At Random (MCAR) using Little’s test, which provided no strong evidence against the MCAR assumption. No clear patterns were observed, suggesting a Not Missing at Random (NMAR) mechanism. Therefore, we used multiple imputation via chained equations to handle missing covariate data, which is appropriate under the MCAR or Missing at Random (MAR) assumptions. The MI model included variables related to missingness and the outcome. Missing data in the dependent variable were not imputed; analyses were restricted to observations with complete outcome data. The significance of the effect size is derived from the beta coefficients of the time variable in both the unadjusted and adjusted models, evaluated at the 95% confidence interval. All the data were analysed using STATA 18.0.

Qualitative data were derived from two sources: written reflections completed by participants and transcripts of yarning sessions conducted with parents and carers. Descriptive narrative analysis was selected as the most appropriate analytic approach for these data, given the exploratory nature of the pilot study and the variation in data form across participant groups.^34^ This approach prioritises the appropriate representation of participants’ voices and experiences and avoids imposing an analytical framework that would exceed what the data can reliably support. It is consistent with Indigenous research methodologies that privilege community knowledge and self-expression as the primary basis for interpretation.^35^

Transcripts and written reflections were independently reviewed by two members of the research team, including a First Nations researcher, to identify recurring ideas and shared experiences regarding program acceptability, perceived benefits, and areas for improvement. This process of independent review followed by consensus discussion supports the credibility and cultural validity of the findings. Areas of interpretive agreement were summarised narratively, and areas of difference were resolved through discussion. Quotes were selected to reflect the range of perspectives expressed by participants, including young people, parents, carers, and Elders, and are presented verbatim when drawn from transcripts and as written by participants when drawn from written reflections, preserving the authenticity of participants’ own words and expression.

### Ethics approval

The University of Queensland’s Human Research Ethics Committee approved this study (2020/HE002899). The study adhered to the AIATSIS Code of Ethics for Aboriginal and Torres Strait Islander Research ^36^ and NHMRC Ethical Conduct in Research with Aboriginal and Torres Strait Islander Peoples and Communities.^37^

## Results

### Participant characteristics

Of 73 adolescents who joined the program, three were excluded from analyses as they did not identify as First Nations, leaving 70 First Nations adolescents who completed baseline data collection. Retention to post-program data collection varied by measurement modality: 38 participants (54.3%) completed the post-program sleep questionnaire, 20 (28.6%) completed the sleep diary (132 nights), and 28 (40.0%) wore the actigraph (Supplementary Figure S1). This differential attrition across modalities should be borne in mind when interpreting cross-modality comparisons, as the subsamples contributing to diary and actigraphy analyses were considerably smaller and may not be representative of the full enrolled cohort.

The sample skewed toward early adolescence, with approximately 73% aged 12–<15 years and was predominantly female (67.1%). The majority identified as Aboriginal only (82.9%), with a further 15.7% identifying as both Aboriginal and Torres Strait Islander. English was the primary home language for 92.8% of participants (Table 2).

**Table 2:** Socio-demographic characteristics of participants at baseline.

|  | **% (n)** |
| --- | --- |
| Total participants: N | 70 |
| Age |  |
| 12-<15 years | 72.9 (51) |
| 15-18 years | 27.1 (19) |
| Sex |  |
| Female | 67.1 (47) |
| Male | 30.0 (21) |
| Missing | 2.9 (2) |
| First Nations status |  |
| Aboriginal | 82.9 (58) |
| Aboriginal and Torres Strait Islander | 15.7 (11) |
| Missing | 1.4 (1) |
| Language spoken at home. |  |
| English | 92.8 (65) |
| home language | 1.4 (01) |
| Missing | 5.7 (4) |
| Post program survey participants | 54.3 (38) |

Contextual data revealed a substantial role of socio-environmental sleep risk factors (Table S1). Although most participants reported access to a safe (88.6%) and clean (87.1%) sleep space, household crowding was common; over half lived in homes with five or more occupants. Approximately 37% resided in noisy or violent neighbourhoods, 37.1% reported going to sleep hungry at least once in the prior week, and 67.1% reported past life experiences that continued to affect their sleep. These figures collectively reflect the broader pattern of socioeconomic and psychosocial challenges, which are an important context to consider when interpreting the sleep health findings.

### Sleep knowledge, sleep health, and psychological distress

Sleep knowledge improved substantially, with the mean composite score increasing from −0.65 (SD 1.23) to 0.82 (SD 1.27), a large effect (Cohen’s *d* = 1.18) that was statistically robust in both unadjusted (β = 1.47, 95% CI: 1.01–1.93, p < 0.001) and adjusted models (β = 1.44, 95% CI: 0.98–1.91, p < 0.001), with minimal attenuation following covariate adjustment. Overall subjective sleep health similarly improved, shifting from −0.82 (SD 1.21) to −0.07 (SD 1.13), a medium-to-large effect (Cohen’s *d* = 0.63) that remained statistically significant after adjustment for age, sex, household occupancy, bedroom access, neighbourhood environment, general health, and going to sleep hungry (adjusted β = 0.68, 95% CI: 0.31–1.04, p < 0.001).

Psychological distress showed a directional reduction in mean composite score from −0.48 (SD 1.81) to −1.09 (SD 2.08), representing a small-to-medium effect (Cohen’s *d* = 0.32) that did not reach statistical significance in unadjusted (β = −0.50, 95% CI: −1.15–0.16, p = 0.134) or adjusted models (β = −0.49, 95% CI: −1.08–0.09, p = 0.097). The wide confidence intervals preclude strong conclusions in either direction, and a modest beneficial effect cannot be excluded.

Pre-to-post regression model results are presented in Figure 1 and Table 3.

**Figure 1:**
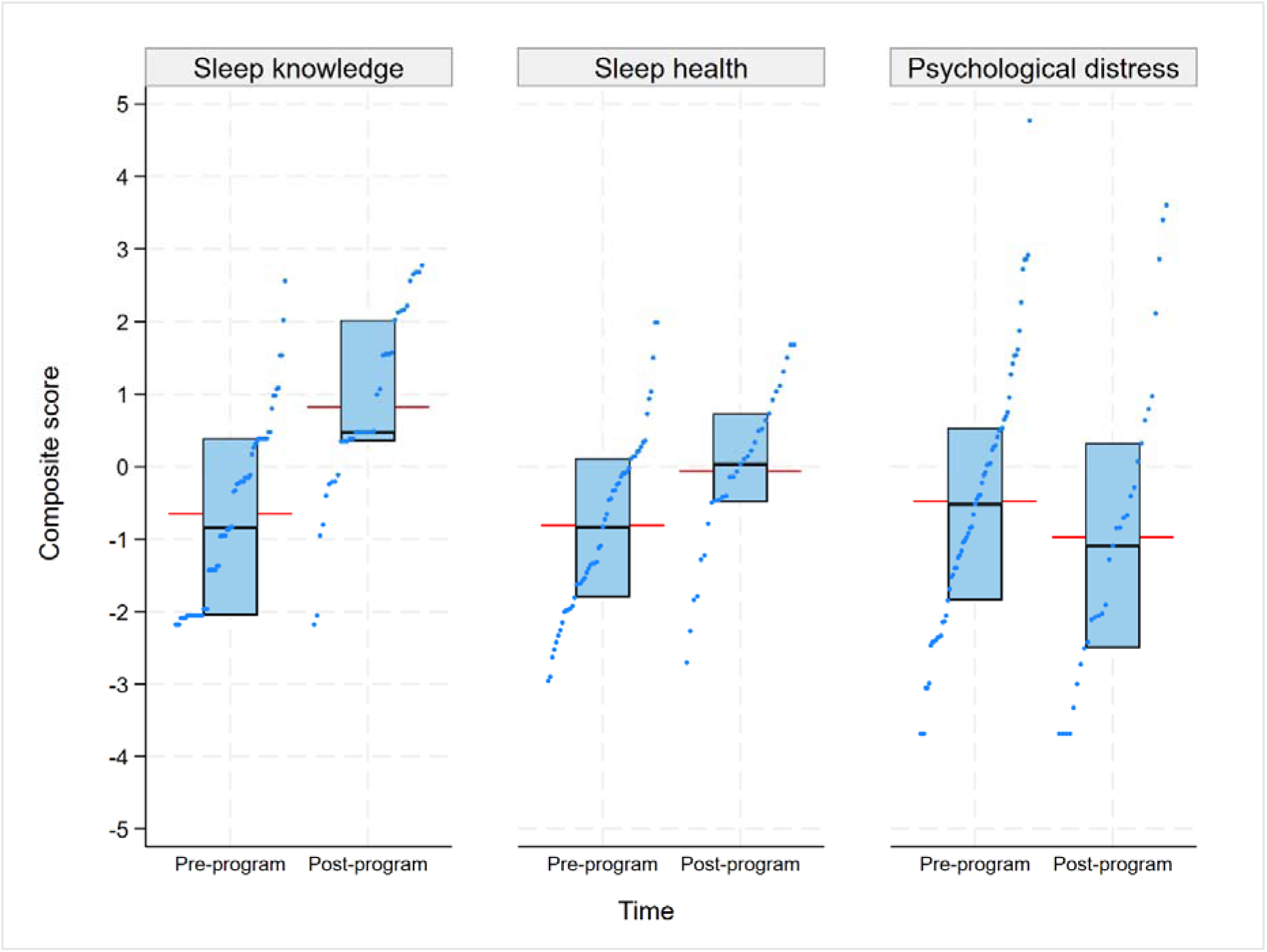
Distribution of participants’ sleep knowledge, sleep health, and psychological distress scores between pre- and post-program surveys.

**Table 3:** Change in sleep knowledge, sleep health, and psychological distress from pre- to post-program.

|  | **N** | **Mean score (SD)** | **Effect size, $\beta$ (95% CI) [P-value]** |  |
| --- | --- | --- | --- | --- |
|  |  |  | **Unadjusted** | **Adjusted\*** |
| Sleep health knowledge | 104 |  |  |  |
| - Pre-program | 67 | -0.65(1.23) |  |  |
| - Post-program | 37 | 0.82(1.27) | **1.47(1.01, 1.93) [<0.001]** | **1.44(0.98, 1.91) [<0.001]** |
| Overall subjective sleep health | 86 |  |  |  |
| - Pre-program | 55 | -0.82(1.21) |  |  |
| - Post-program | 31 | -0.07(1.13) | **0.75(0.38, 1.12) [<0.001]** | **0.68(0.31, 1.04) [<0.001]** |
| Psychological distress | 96 |  |  |  |
| - Pre-program | 65 | -0.48(1.81) |  |  |
| - Post-program | 31 | -1.09(2.08) | -0.50(-1.15, 0.16) [0.134] | -0.49(-1.08, 0.09) [0.097] |

\*P<20 in bivariate analysis were included in the adjusted models. Covariates for the sleep health knowledge model: age, sex, and language spoken at home. Covariates for the overall sleep health, subjective and objective sleep timing and continuity, and subjective sleep sufficiency models: age, sex, number of people at home, having one's own room, neighbourhood condition, general health status, and going to sleep hungry. Covariates for the psychological distress model: age, sex, neighbourhood condition, general health status, and going to sleep hungry.

### Individual sleep parameters

Detailed findings across measurement modalities are presented in (Supplementary table 2). The overarching pattern was consistent directional improvement across subjective measures, with no statistically significant change in objective sleep parameters.

A striking cross-modality discrepancy in baseline sleep duration estimates warrants acknowledgement. Only 12.8% of participants met the age-recommended sleep duration (≥9 hours) by actigraphy, compared with 57.1% by sleep diary and 78.1% by questionnaire. Across subjective measures, directional improvements were observed in sleep timing, quality, onset latency, and duration, though most did not reach statistical significance, likely reflecting limited power rather than the absence of an effect.

The most notable exception was questionnaire-reported wake time, which advanced significantly by approximately one hour (p = 0.018). Sleep diary-recorded duration showed a near-significant increase of approximately 36 minutes (8.2 to 8.8 hours, p = 0.065), and the proportion rating sleep quality as “great” increased from 31.3% to 50.0%, though this did not reach significance (p = 0.204). Two functionally important outcomes approached significance: sleep affecting daytime functioning declined from 62.3% to 41.7% (p = 0.057), and reported sleep concerns declined from 50.0% to 36.8% (p = 0.077).

Regarding sleep regularity, sleep diary-derived bedtime variability (mean within-person standard deviation across nights) declined from 73.0 to 65.0 minutes and wake time variability from 66.0 to 58.2 minutes post-program, indicating marginally more consistent sleep timing, though neither change was statistically significant (p = 0.894 and p = 0.748).

### Program acceptability

Young people and their parents and carers (n = 18) participated in yarning sessions at program completion to share their experiences, perceived benefits, and views on the program. Feedback was collected through yarning sessions facilitated by First Nations Sleep Coaches in culturally safe community settings.

Participants described the program as engaging, relevant, and enjoyable, emphasising its variety and interactive design. As one participant explained:

*”I liked it because we don’t do the same thing every day. We do a whole variety of things, and it is very entertaining. With this program, I have been able to get a better sleep with more hours.”*

Another reflected:

*I liked all the different activities; there was nothing I didn’t like*.

Parents noted the practical benefits of the sessions, particularly those focused on sleep hygiene and relaxation:

*”The learnings in the sessions were beneficial, especially the ones around the healthy sleep hygiene and the relaxation, which would help create a calming, relaxed setting to promote a quality sleep.”*

Young participants also described concrete changes in their own sleep behaviours, including earlier bedtimes and more consistent routines:

*”I have had changes in my sleep such as going to bed earlier. I don’t stay up as long anymore and usually go to bed before 10–11. My sleep has been getting better.”*

Session rating data supported these qualitative accounts. Across all sessions, participants consistently rated the program highly on engagement, delivery, and content (n = 194 ratings). Ratings of “Deadly” (Excellent), the highest rating on the culturally adapted scale, increased steadily over time, peaking at 75% in Session 5, while “Needs Improvement” remained below 7% throughout. Participants highlighted hands-on interactive activities, including Walk on Country, as key contributors to their experience.

Community Elders expressed strong endorsement of the program, framing it as an important step in strengthening intergenerational health knowledge and supporting early intervention. One Elder shared:

*”This program is there letting everyone else know that things are happening to our younger generations out there, and this study is one of the best things that could happen to let us know why the children are not getting enough sleep.”*

Another reflected:

*”I think the biggest part is we’re catching our kids. You know, we’re catching our young ones before, because I think by the time they get to parenthood, they’ll be able to help their children and guide their children to a better life, because sleep is most important for everybody.”*

Overall, the program was acceptable to First Nations adolescents and their families, with strong and sustained engagement across sessions and clear community endorsement from Elders.

## Discussion

This study shows that a sleep health program, developed in collaboration with First Nations communities, was both welcomed and offers promising results to improve sleep health of First Nations young people. Our results suggest improvements in sleep health and a possible reduction in psychological distress. What made the program work was its interactive, strengths-based design, its grounding in First Nations knowledge, and being led by trusted community members. These results highlight the importance of approaches guided by community voices in supporting sleep health in First Nations settings. Every community has its own needs and strengths, so the results from one community may not fit everywhere. However, the co-design journey has offered practical tools and new ways of working to guide sleep health equity efforts in other communities.

The large effect sizes observed for sleep health knowledge are encouraging and indicate that the program successfully translated attendance into meaningful learning. It is well established that participant knowledge and intention are not necessarily sufficient to enact behaviour change (the intention-behaviour gap) ^38^, and this study’s findings reflect that complexity. Importantly, the absence of statistically significant change in some sleep measures should not be read as evidence that the program did not work. Participants themselves described feeling better rested, more aware of their sleep, and more settled in their routines, and these improvements are meaningful outcomes in their own right, particularly in a setting where sleep health is challenging to prioritise and protect

Where changes were observed, they were consistent and coherent. Self-reported sleep health improved significantly, with earlier bedtimes, reduced short sleep duration, improved sleep quality, and reduced daytime sleepiness. The absence of significant change in objective actigraphy measures is not unexpected and does not contradict the self-reported findings. It is well documented that self-reported and objectively measured sleep frequently diverge, and this study is no exception. ^39^ It is also worth considering whether the two-week follow-up interval after program completion was sufficient to detect sustained physiological change, or whether a longer follow-up would reveal a greater or lesser effect. Future studies with extended follow-up periods would help resolve this question.

The sustained and growing engagement across all five sessions highlights the program’s acceptability. The progressive improvement in session ratings, with the highest scores recorded in the final sessions, suggests that the yarning-based format, the relationships built with Sleep Coaches, and the hands-on activities became more meaningful to participants as the program unfolded, rather than losing momentum over time. This pattern of increasing rather than declining engagement over a ten-week program is notable, particularly given the challenges of retaining adolescents in community-based health initiatives in remote settings.

Qualitative feedback deepened this picture. Participants described the program as enjoyable, varied, and directly connected to their lives, not an external health message delivered to them, but something they felt part of. Parents valued the practical focus on sleep hygiene and relaxation, and young people described real changes in their own bedtime routines. The fact that participants could point to concrete behavioural shifts going to bed earlier, sleeping more consistently and connect those shifts to what they learned in the program suggests the content landed in a way that mattered beyond the sessions themselves.

Community Elders’ endorsement of the program carries significant weight. Elders’ advocacy for extending the program to other First Nations communities reflects genuine confidence in its value, appropriateness, and fit with community priorities. This kind of community-level endorsement is arguably as important an indicator of program success as any quantitative outcome.

This program offered valuable insights into the realities of delivering community-led health initiatives in remote First Nations settings. Environmental disruptions such as severe weather events and bushfire risks occasionally delayed travel and field activities. These external factors extended project timelines and necessitated flexibility in delivery schedules. Like many other remote communities, high workforce turnover posed an additional challenge, requiring repeated recruitment, training, and mentoring to sustain program continuity. Engaging young participants also proved challenging. Many adolescents in the target group were disengaged from school and had irregular sleep-wake patterns, making it hard to ensure consistent participation. For some, the 10-week after-school sessions clashed with other commitments. As a remote community with a mobile population, several families relocated for work or personal reasons during the study, resulting in many young people leaving with their families before program completion. Despite these challenges, the program maintained strong engagement throughout, and First Nations Sleep Coaches continued supporting participants beyond the research project’s duration.

Several factors were central to this program’s success. Privileging First Nations voices and blending traditional and Western sleep knowledge created a foundation of mutual respect and shared decision-making. The Elders Steering Group ensured cultural values guided all aspects of the program. Collaboration with local artists, radio stations, and First Nations-owned businesses extended the program’s reach and reinforced community ownership. Employing, training, and mentoring Aboriginal Youth Workers as Sleep Coaches was perhaps the most critical structural decision; it ensured the program was delivered by and for the community, built local capacity that outlasted the project, and created trusted relationships that no external team could replicate. Pausing activities during culturally significant periods, including Sorry Business (the customary mourning, ceremonies, and obligations observed by many Aboriginal communities that follow the death of a community member), demonstrated genuine respect for community protocols and strengthened trust.

This study has several limitations that should be acknowledged. The absence of a control group means observed changes cannot be attributed solely to the program, and the pre–post design cannot rule out external influences on outcomes. Future research should include a control or comparison group to strengthen causal inferences. Attrition occurred at two distinct stages. First, ten young people enrolled and completed baseline data collection but did not commence the program: nine owing to family relocation and one for personal reasons. Among those who began the program, engagement remained high. However, post-program data collection proved challenging, as several participants did not attend follow-up assessment sessions. This loss to follow-up reduced the analytic sample, potentially limiting statistical power and introducing bias. Funding constraints precluded a formal long-term follow-up, leaving the sustainability of behaviour change unconfirmed. Funding constraints precluded a formal long-term follow-up, leaving the sustainability of behaviour change unconfirmed. While the program was designed from the outset with cultural safety as a guiding principle through co-design with Elders, community-led delivery by Aboriginal Youth Workers, and integration of First Nations pedagogical practices, formal measurement of cultural safety was beyond the scope of this study. Future research should develop specific, community-validated indicators to assess cultural safety as an outcome in its own right, rather than treating it as an assumed feature of co-designed programs.

The lessons from this project point clearly toward embedding culturally adapted, community-led sleep health promotion within broader health and education systems. Integrating sleep health into primary care, youth well-being services, and, where appropriate, school-based programs, guided by First Nations leadership, offers a promising pathway toward sustainable change and reduced sleep health inequities for First Nations young people.

## Conclusion

This project demonstrated that a culturally grounded, community-led approach to sleep health can meaningfully engage First Nations adolescents, families, and communities in addressing an important yet under-recognised determinant of well-being. The co-design process, driven by Elders, young people, parents, and service providers, ensured that the program reflected community values, priorities, and lived experiences, leading to high satisfaction and strong local ownership. We have demonstrated that such an intervention can improve knowledge of sleep health, healthy sleep behaviours, and self-reported sleep duration. Notably, the outcomes highlight that sleep health interventions are most effective when rooted in First Nations worldviews and co-designed in partnership with First Nations communities. The findings provide new evidence in the field of First Nations adolescent sleep health and demonstrate a scalable model that can inform future community-based sleep health initiatives.

## Data Availability

The study protocol is available elsewhere. The study tools are available on request. Individual participant data is not made available to external parties

## Acknowledgement

We extend our deepest gratitude to the First Nations community members of Mount Isa, particularly Kalkadoon Elders, whose invaluable insights, support, and collaboration guided this project. We also thank Mithangkaya Nguli Young People Ahead Youth & Community Services Indigenous Corporation ICN 9842 team for their support and assistance. This work would not have been possible without their generous sharing of time, resources, and expertise.

## Contributors

Yaqoot Fatima: Conceptualisation, Funding acquisition, Methodology, Project administration, Aanlysis, Supervision, Writing—original draft, Writing—review & editing

Roslyn Von Senden: Methodology, Project administration, Supervision, Writing—review & editing

M Mamun Huda: Methodology, Analysis, Writing—original draft, Writing—review & editing

Aunty Joan Marshall: Cultural mentoring, Conceptualisation, Methodology, Writing—review

Daniel Sullivan: Project administration, Analysis, Writing—original draft, Writing—review & editing

Romola Bucks: Conceptualisation, Funding acquisition, Methodology, Writing—review & editing

Azhar Hussain Potia: Methodology, Writing—original draft, Writing—review & editing

Simon Smith: Funding acquisition, Methodology, Writing—review & editing

Sarah Blunden: Funding acquisition, Methodology, Writing—review & editing

Lisa McDaid: Funding acquisition, Methodology, Writing—review & editing

Markesh Fanti: Methodology, Project administration, Writing—review & editing

Peter Eastwood: Funding acquisition, Methodology, Writing—review & editing

Stephanie Yiallourou: Funding acquisition, Methodology, Writing—review & editing

Jennifer Walsh: Resources, Methodology, Writing—review & editing

Abdullah Mamun: Funding acquisition, Methodology, Writing—review & editing

Stephanie King: Funding acquisition, Methodology, Writing—review & editing

Sharon Varela: Funding acquisition, Methodology, Writing—review & editing

Shaun Solomon: Funding acquisition, Methodology, Writing—review & editing

Timothy Skinner: Conceptualisation, Funding acquisition, Methodology, Writing—original draft, Writing—review & editing

## Data sharing statement

The study protocol is available elsewhere ^22^. The study tools are available on request. Individual participant data is not made available to external parties.

## Supplementary Figures and Tables

**Figure S1:**
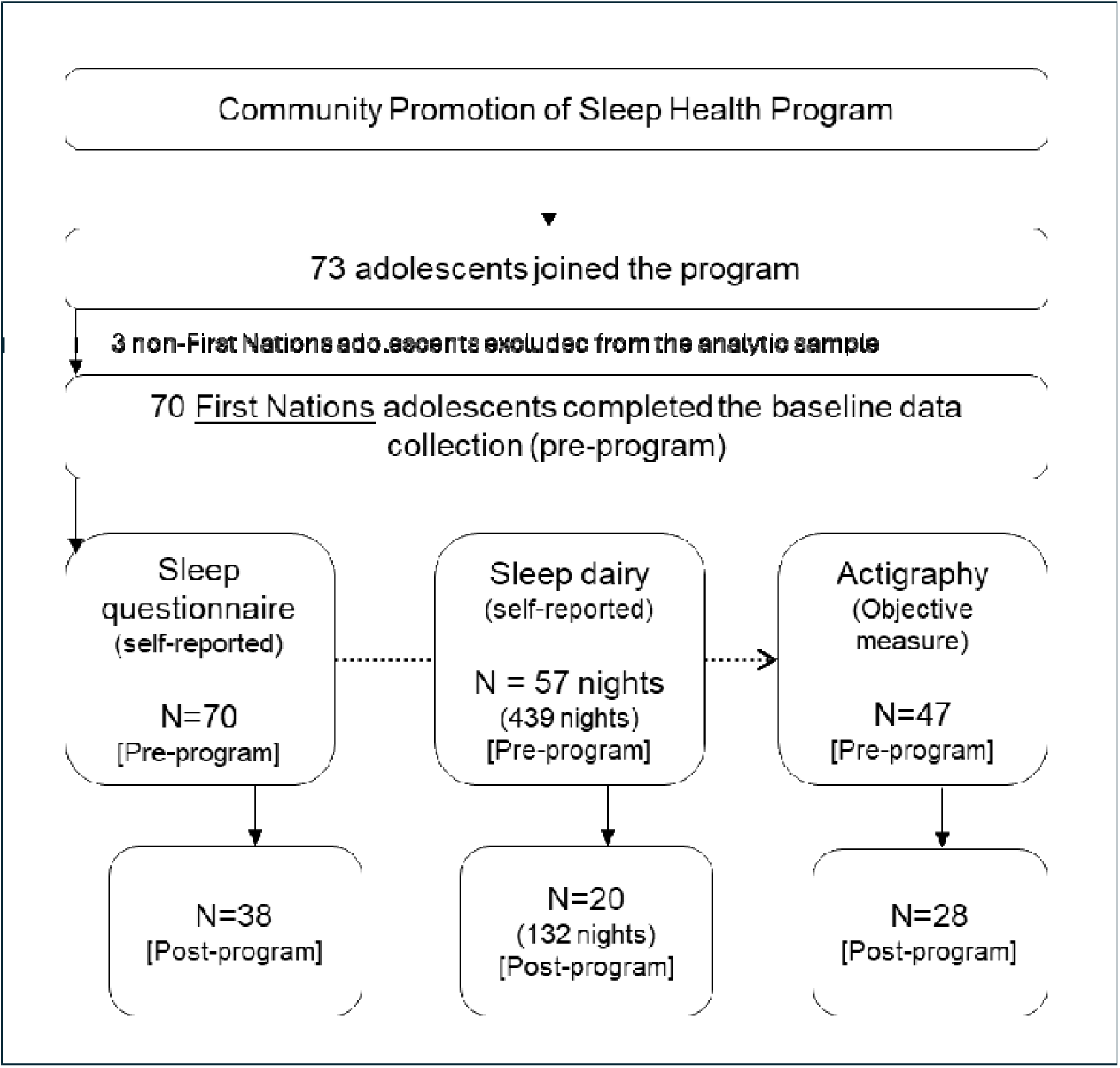
Overview of the analytical sample section.

**Table S1:** Distribution of participants’ sleep environment and behavioural variables.

|  | **% (n)** |
| --- | --- |
| **Sleep environment** |  |
| **Number of people at home** |  |
| <5 | 42.9(30) |
| 5-9 | 34.3(24) |
| 10+ | 18.6(13) |
| Missing | 4.3(3) |
| **Number of rooms** |  |
| <3 | 45.7(32) |
| 3-4 | 52.9(37) |
| Missing | 1.4(1) |
| **Had own room** |  |
| No | 30.0(21) |
| Yes | 68.6(48) |
| Missing | 1.4(1) |
| **Safe sleeping space** |  |
| No or not always | 8.6(6) |
| Yes | 88.6(62) |
| Missing | 2.9(2) |
| **Clean sleeping space** |  |
| No or not always | 11.4(8) |
| Yes | 87.1(61) |
| Missing | 1.4(1) |
| **Neighbourhood** |  |
| Noisy and violent | 10.0(7) |
| Noisy | 27.1(19) |
| Quiet or safe | 62.9(44) |
| **General health** |  |
| Good | 68.6(48) |
| Okay | 30.0(21) |
| Missing | 1.4(1) |
| **Went to sleep hungry in the past week** |  |
| None or <1 time | 61.4(43) |
| >=1 time | 37.1(26) |
|  | **% (n)** |
| Missing | 1.4(1) |
| **Life events affecting sleep** |  |
| No | 22.9(16) |
| Yes | 67.1(47) |
| Missing | 10.0(7) |
| **Behavioural factors** |  |
| **Screen time (per day/prior to bed)** |  |
| <2 hours | 17.1(12) |
| 2-4 hours | 32.9(23) |
| 4+ hours | 47.1(33) |
| Missing | 2.9(2) |
| **Caffeinated drinks (per day)** |  |
| None | 57.1(40) |
| Once | 24.3(17) |
| More than once | 7.1(5) |
| Missing | 11.4(8) |
| **Frequency of $\geq 20$ minutes of physical activity (times/week)** |  |
| None | 11.4(8) |
| <1 | 14.3(10) |
| >1 | 71.4(50) |
| Missing | 2.9(2) |
| **Homework or school assignments affect sleep** |  |
| No | 38.6(27) |
| Sometimes | 31.4(22) |
| Yes | 22.9(16) |
| Missing | 7.1(5) |

**Table S2:**
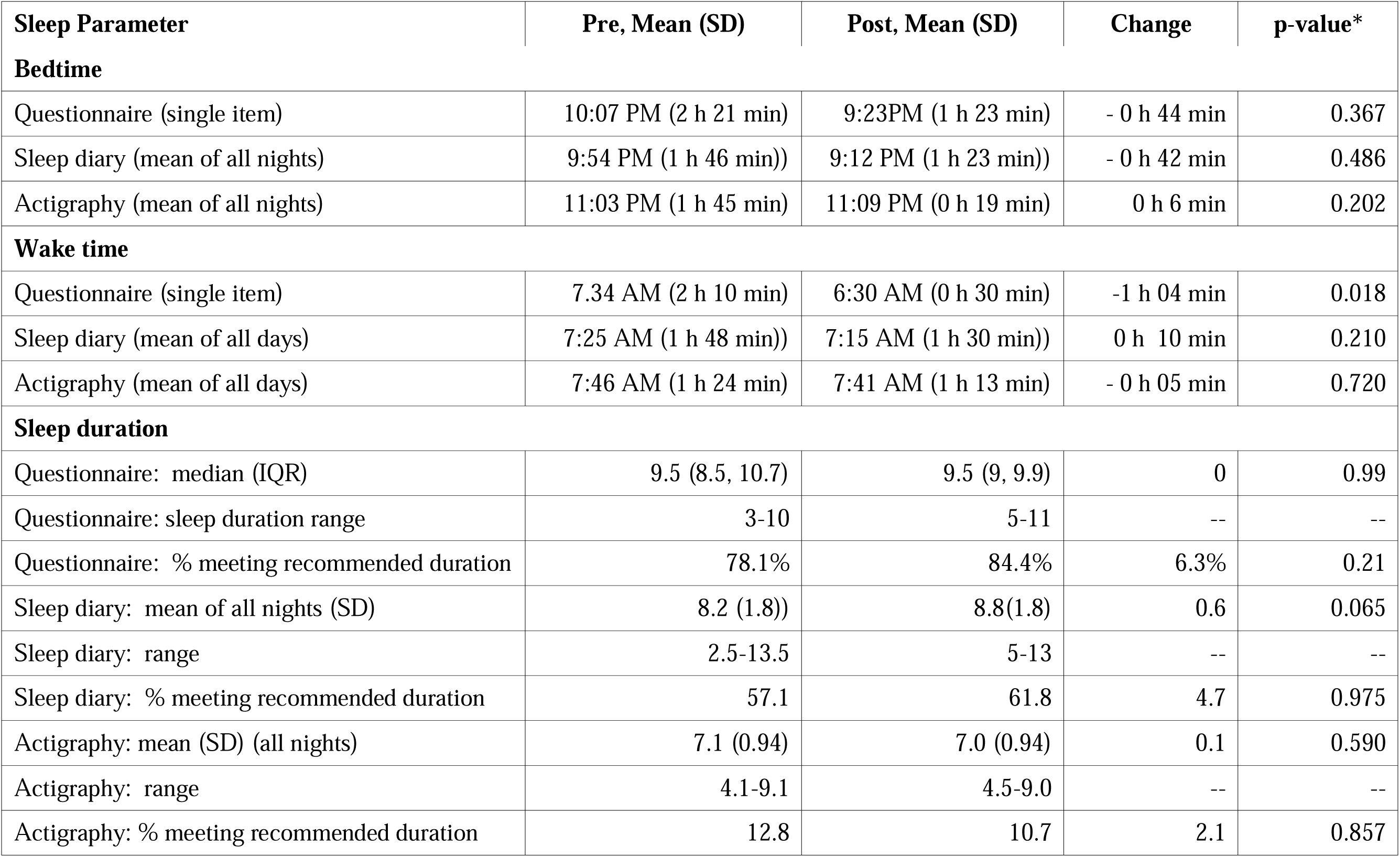
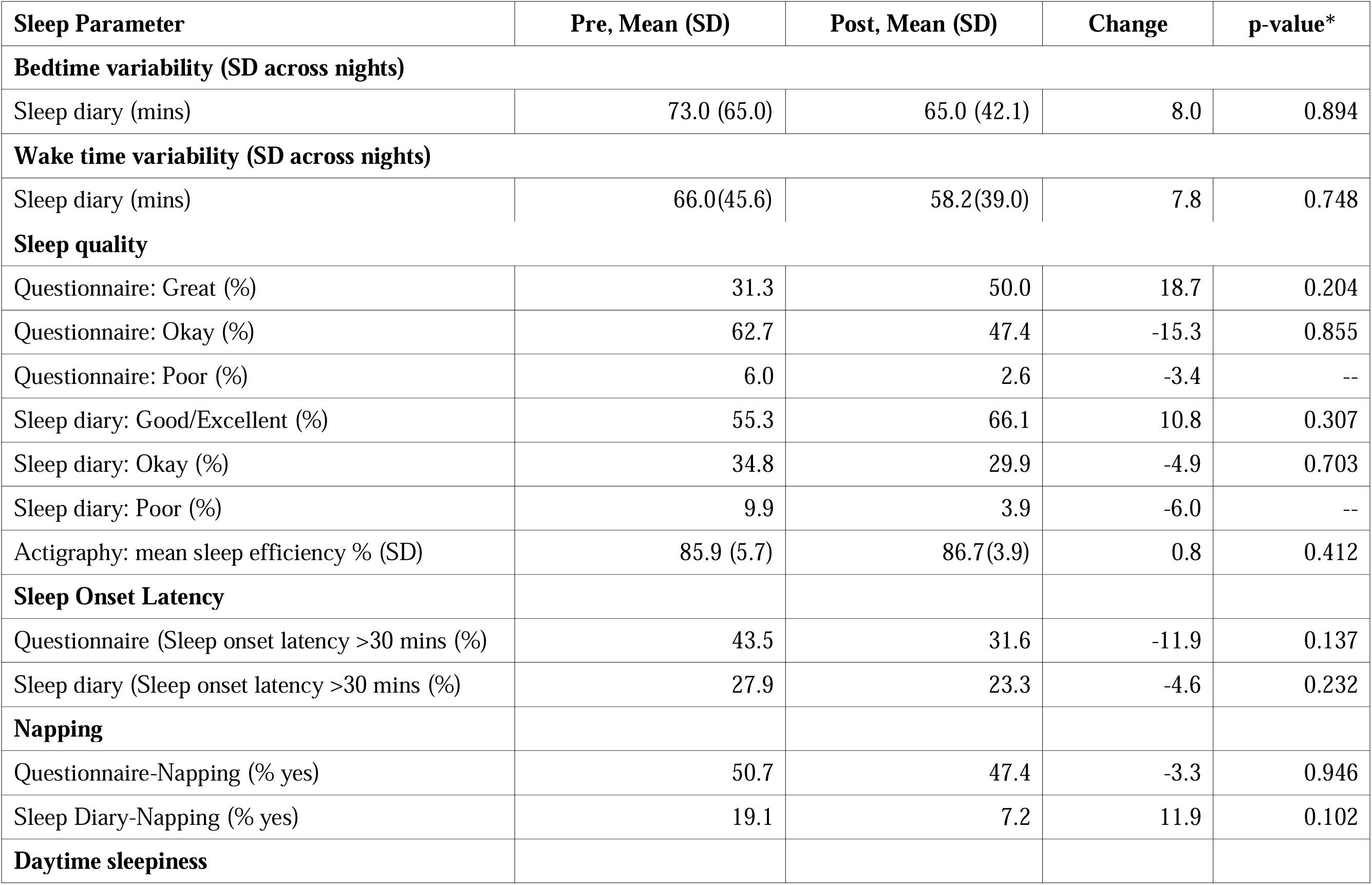
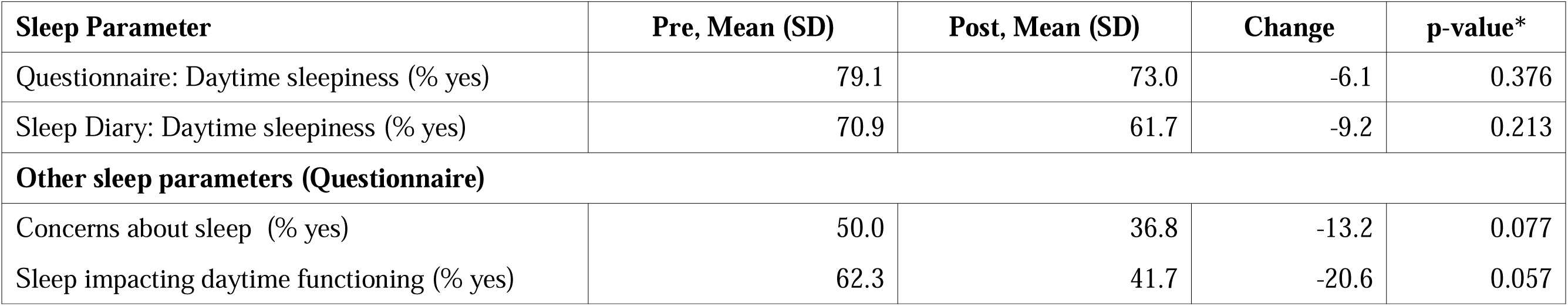
Pre- and post-program distribution of individual sleep parameters from self-reported questionnaire, sleep diary (seven nights) and actigraphy (seven nights)

| **Sleep Parameter** | **Pre, Mean (SD)** | **Post, Mean (SD)** | **Change** | **p-value\*** |
| --- | --- | --- | --- | --- |
| **Bedtime** |  |  |  |  |
| Questionnaire (single item) | 10:07 PM (2 h 21 min) | 9:23PM (1 h 23 min) | - 0 h 44 min | 0.367 |
| Sleep diary (mean of all nights) | 9:54 PM (1 h 46 min)) | 9:12 PM (1 h 23 min)) | - 0 h 42 min | 0.486 |
| Actigraphy (mean of all nights) | 11:03 PM (1 h 45 min) | 11:09 PM (0 h 19 min) | 0 h 6 min | 0.202 |
| **Wake time** |  |  |  |  |
| Questionnaire (single item) | 7.34 AM (2 h 10 min) | 6:30 AM (0 h 30 min) | -1 h 04 min | 0.018 |
| Sleep diary (mean of all days) | 7:25 AM (1 h 48 min)) | 7:15 AM (1 h 30 min)) | 0 h 10 min | 0.210 |
| Actigraphy (mean of all days) | 7:46 AM (1 h 24 min) | 7:41 AM (1 h 13 min) | - 0 h 05 min | 0.720 |
| **Sleep duration** |  |  |  |  |
| Questionnaire: median (IQR) | 9.5 (8.5, 10.7) | 9.5 (9, 9.9) | 0 | 0.99 |
| Questionnaire: sleep duration range | 3-10 | 5-11 | -- | -- |
| Questionnaire: % meeting recommended duration | 78.1% | 84.4% | 6.3% | 0.21 |
| Sleep diary: mean of all nights (SD) | 8.2 (1.8)) | 8.8(1.8) | 0.6 | 0.065 |
| Sleep diary: range | 2.5-13.5 | 5-13 | -- | -- |
| Sleep diary: % meeting recommended duration | 57.1 | 61.8 | 4.7 | 0.975 |
| Actigraphy: mean (SD) (all nights) | 7.1 (0.94) | 7.0 (0.94) | 0.1 | 0.590 |
| Actigraphy: range | 4.1-9.1 | 4.5-9.0 | -- | -- |
| Actigraphy: % meeting recommended duration | 12.8 | 10.7 | 2.1 | 0.857 |

| Sleep Parameter | Pre, Mean (SD) | Post, Mean (SD) | Change | p-value* |
| --- | --- | --- | --- | --- |
| **Bedtime variability (SD across nights)** |  |  |  |  |
| Sleep diary (mins) | 73.0 (65.0) | 65.0 (42.1) | 8.0 | 0.894 |
| **Wake time variability (SD across nights)** |  |  |  |  |
| Sleep diary (mins) | 66.0(45.6) | 58.2(39.0) | 7.8 | 0.748 |
| **Sleep quality** |  |  |  |  |
| Questionnaire: Great (%) | 31.3 | 50.0 | 18.7 | 0.204 |
| Questionnaire: Okay (%) | 62.7 | 47.4 | -15.3 | 0.855 |
| Questionnaire: Poor (%) | 6.0 | 2.6 | -3.4 | -- |
| Sleep diary: Good/Excellent (%) | 55.3 | 66.1 | 10.8 | 0.307 |
| Sleep diary: Okay (%) | 34.8 | 29.9 | -4.9 | 0.703 |
| Sleep diary: Poor (%) | 9.9 | 3.9 | -6.0 | -- |
| Actigraphy: mean sleep efficiency % (SD) | 85.9 (5.7) | 86.7(3.9) | 0.8 | 0.412 |
| **Sleep Onset Latency** |  |  |  |  |
| Questionnaire (Sleep onset latency >30 mins (%)) | 43.5 | 31.6 | -11.9 | 0.137 |
| Sleep diary (Sleep onset latency >30 mins (%)) | 27.9 | 23.3 | -4.6 | 0.232 |
| **Napping** |  |  |  |  |
| Questionnaire-Napping (% yes) | 50.7 | 47.4 | -3.3 | 0.946 |
| Sleep Diary-Napping (% yes) | 19.1 | 7.2 | 11.9 | 0.102 |
| **Daytime sleepiness** |  |  |  |  |
| Questionnaire: Daytime sleepiness (% yes) | 79.1 | 73.0 | -6.1 | 0.376 |
| Sleep Diary: Daytime sleepiness (% yes) | 70.9 | 61.7 | -9.2 | 0.213 |
| **Other sleep parameters (Questionnaire)** |  |  |  |  |
| Concerns about sleep (% yes) | 50.0 | 36.8 | -13.2 | 0.077 |
| Sleep impacting daytime functioning (% yes) | 62.3 | 41.7 | -20.6 | 0.057 |

